# MAP-DyS: An Interactive Framework for Mapping Analytic Decision Pathways in Subtyping Research

**DOI:** 10.1101/2025.07.23.25332032

**Authors:** Anna Yi Leung, Daniel Kristanto, Carsten Gießing, John PA Ioannidis, Andrea Hildebrandt, Xenia Schmalz

**Author notes:** Corresponding author: Anna Yi Leung. Department of Child and Adolescent Psychiatry, Psychosomatics, and Psychotherapy, University Hospital, Ludwig-Maximilians-University of Munich, Nussbaumstrasse 5a, 80336 Munich, Germany. Shared first authorship (Anna Yi Leung, Daniel Kristanto). Shared senior authorship (Andrea Hildebrandt, Xenia Schmalz). In alphabetical order (Carsten Gießing, John PA Ioannidis, Andrea Hildebrandt, Xenia Schmalz).

## Abstract

Subtyping approaches are widely used in psychological and cognitive research to classify individuals into subgroups based on shared behavioural or cognitive characteristics, particularly when addressing heterogeneity in developmental conditions such as developmental dyslexia. However, subtyping analyses involve numerous methodological decisions, including the choice of theoretical models, data preprocessing strategies, performance indices, and statistical techniques. Variability in these analytic decisions can lead to inconsistent subgroup identification and complicate comparisons across studies, yet such methodological variability is rarely systematically mapped. To support transparent examination of analytic decision pathways in subtyping research, we developed MAP-DyS, an open-source interactive Shiny app that enables researchers to explore and compare methodological choices across subtyping studies. To demonstrate the approach, we compiled a corpus of developmental dyslexia subtyping studies identified through a systematic search of four academic databases and extracted key analytic decision points reported in these studies. The resulting dataset illustrates substantial variability in theoretical models, preprocessing procedures, statistical methods, and reported subgroups. MAP-DyS allows users to interactively visualise these decision pathways and examine how different methodological configurations are represented across the literature. By making analytic variability transparent and navigable, the tool supports researchers in critically evaluating existing subtyping practices and designing more transparent and reproducible subtyping studies. Although demonstrated using developmental dyslexia research, the framework is designed to be adaptable to other areas of psychological and behavioural research that employ subtyping or classification approaches.

## Introduction

Subtyping is widely used across psychology, cognitive science, and biomedicine to characterise heterogeneity within populations by grouping individuals into distinct profiles based on cognitive, behavioural, or clinical measures (e.g., Feczko et al., 2019; Richards & Hewstone, 2001). These subgroup classifications are often interpreted as reflecting meaningful underlying mechanisms and are used to inform theory development, diagnosis, and intervention (Benkarim et al., 2022; Gearin et al., 2022; Lorusso et al., 2011; Wolf et al., 2024). However, subtyping results are highly sensitive to analytic decisions (Brusco et al., 2017; Burns et al., 2023; Sorgente et al., 2025; Yuan et al., 2023). In practice, subtyping outcomes are shaped by a multistage analytic pipeline that includes decisions about theoretical frameworks (e.g., Hooper & Willis, 2013), data preprocessing strategies (e.g., transformation and standardisation; see also Acedo-Terrades et al., 2025), and statistical techniques (e.g., cluster analysis, latent class analysis, quantile classification; see relevant cases: Alosh et al., 2015; Horne et al., 2020). These decisions define the analytic space within which subtyping approaches can be implemented. As a result, studies addressing similar research questions may arrive at qualitatively different subgroup structures, raising questions about the robustness and interpretability of resulting classifications.

This challenge reflects a broader issue of researcher degrees of freedom in complex analytic workflows. While some authors have called for standardisation in subtyping approaches (Zhao et al., 2019), a single, widely accepted and uncontested approach may be utopian to expect, given the conceptual and methodological diversity of the field. Multiverse analysis offers an alternative method when there is no consensus on standardisation by systematically mapping and evaluating justifiable analytic decisions and the resulting outcomes (see comprehensive reviews on multiverse analysis: Patel et al., 2015; Short et al., 2025; Steegen et al., 2016). This approach has been successfully applied in fields such as clinical psychology, social psychology, and sociology to quantify how arbitrary analytic decisions impact results. A critical prerequisite for applying a multiverse perspective is the explicit identification of the underlying decision space.

However, in subtyping research, this decision space is rarely systematically defined. To evaluate whether a result is robust across analytic choices, one must first identify which decisions are available, commonly used, and theoretically or methodologically justified (Short et al., 2025). Without a systematic map of these researcher degrees of freedom—ranging from broad theoretical priors to granular preprocessing steps—it is difficult to design an informed multiverse analysis or to interpret variability across studies. This limitation highlights the need for a systematic framework that maps the landscape of analytic decision pathways.

To address this gap, we introduce MAP-DyS (“Mapping Dyslexia Subtypes”), an open-source, interactive framework with a Shiny app for mapping analytic decision pathways in subtyping research. MAP-DyS operates at the level of published academic articles, visualising a dataset that captures the full range of analytic decisions reported across empirical studies. We demonstrate this framework using developmental dyslexia as a use case. Developmental dyslexia (hereinafter referred to as dyslexia) is a neurodevelopmental disorder characterised by persistent difficulties in reading and spelling, despite adequate intelligence and educational opportunities (see reviews: Snowling et al., 2020; Wolf et al., 2024), affecting approximately 5% to 20% of individuals depending on assessment methods and populations (Wagner et al., 2020; Yang et al., 2022). Dyslexia has long been recognised as a heterogeneous condition, with variability in both underlying mechanisms and cognitive profiles (Friedmann & Coltheart, 2018; Hanley, 2017; Peterson & Pennington, 2012; Snowling, 2008; Wolf et al., 2024). This heterogeneity includes variability in both definition and causal mechanisms, as well as the individual differences in the cognitive profiles of dyslexic readers (Elliott & Grigorenko, 2014; Friedmann & Coltheart, 2018; Snowling, 2008; Snowling et al., 2020; Wolf et al., 2024), such as differences in phonological processing (Navas et al., 2014; Snowling et al., 2020), rapid automatised naming (RAN; e.g., Araújo & Faísca, 2019; Siddaiah & Padakannaya, 2015), and working memory (de Assis Leão et al., 2023; Smith-Spark, 2018), among others (see relevant reviews, e.g., Elliott & Grigorenko, 2014, 2024; Wolf et al., 2024). Subtyping approaches have therefore been used to operationalise this heterogeneity by identifying groups of individuals with distinct profiles. At the same time, dyslexia subtyping research exemplifies the methodological variability described above, making it a suitable case for demonstrating how analytic decisions shape subtyping outcomes.

Rather than prescribing a single optimal approach, MAP-DyS enables researchers to interrogate how different analytic pathways shape subtyping results, assess the comparability of findings across studies, and identify areas where methodological variability may influence interpretation. In this sense, our framework shifts the focus from identifying subtypes to understanding how subtypes are constructed. More broadly, the framework contributes to improving transparency, supporting reproducibility, and facilitating more informed research design in subtyping studies across psychological and behavioural science. The Shiny app, along with all relevant R scripts and datasets, is publicly available at https://github.com/kristantodan12/Dyslexia_subtyping.

## Corpus Construction and Coding Framework

### Systematic Literature Search

Following the “Preferred Reporting Items for Systematic Review and Meta-Analysis (PRISMA)” for protocols (Moher et al., 2015), we conducted a systematic literature search to identify studies that used methods of subtyping readers with developmental dyslexia. The protocol for this literature search was not pre-registered. We performed the search across four bibliographic databases. The primary databases included the Excerpta Medica dataBASE (Embase, which includes the MEDLINE and PubMed databases), APA PsycInfo, and Education Resources Information Centre (ERIC), where the latter two database searches were conducted via the search engine, EBSCOhost; the supplementary database included OSF Preprints, which covered grey literature or unpublished literature. We selected these databases according to their broad coverage and search efficiency (Bramer et al., 2017), ensuring representation across key fields in which dyslexia subtyping studies are published, including psychology, behavioural sciences, education, and biomedicine. There were no restrictions on the publication year. The first author conducted the literature search in December 2022, forming the basis for the present implementation of the MAP-DyS framework. The search string used on Embase and EBSCOhost was (dyslex* OR “poor reader*” OR “reading disorder”) AND (subtype* OR subgroup* OR profil*). On EBSCOhost, we selected items that had linked full texts and conducted the search within all texts, including the full texts, titles, abstracts, and keywords. On OSF Preprints, we used the same search string as on Embase and EBSCOhost, using the default settings, except that truncation was not used to optimise the search performance (see Bramer et al., 2017, for a detailed comparison of suitable academic search engines for systematic reviews).

### Eligibility Criteria

A record was eligible if it reported: (1) the results of at least one original empirical study and (2) the identification of the cognitive and behavioural profiles/subtypes/subgroups of readers with reading difficulty. If a study reported the subtypes of a particular cognitive ability (e.g., seeking subtypes of executive functions of readers) but not the types of dyslexic readers, such a record was excluded. We also excluded studies that used only neural measures (e.g., fMRI) without any behavioural measures to subtype readers. However, we included records that used behavioural measures alongside neurophysiological measures (e.g., visual search task + EEG) for subtyping. In addition, we excluded studies that examined subtypes based solely on genetic data (e.g., genome-wide association studies), as the focus of this study was on cognitive and behavioural subtyping.

It is worth noting that readers with reading difficulty are sometimes referred to using different terminology, such as “poor readers”, “readers with reading difficulty”, “readers with reading disabilities”, “dyslexics”, and “readers with developmental dyslexia”, etc. If poor readers named in any of the above terms were included in the subtype identification process, such a study still fulfilled the second eligibility criterion. Also, we included studies in which the poor readers in the reported sample were comorbid with other disorders, such as Attention-Deficit Hyperactivity Disorder (ADHD), dyscalculia, and executive functioning disorder. However, studies were excluded if the sample did not include individuals with developmental dyslexia, such as those consisting solely of typically developing readers or individuals with unrelated language difficulties.

### Screening Process

Two raters (Rater 1 and Rater 2) conducted the screening using the R Shiny app, “revtools” (Westgate, 2019). The first author (Rater 1) screened all records. Rater 2 screened a randomly selected 20% of the items to assess inter-rater reliability. This subset was blind-coded. We computed the inter-rater reliability of the 20% of records using Cohen’s kappa (κ). The two raters resolved any discrepancies through discussion until consensus was reached. If κ was ≥ .7, the agreed-upon 20% and the remaining 80%, only screened by Rater 1, were retained as the final dataset. If κ was below .7, both raters jointly examined the reasons for disagreement, after which Rater 1 re-reviewed the remaining 80% to finalise the dataset for the next stage of data extraction.

The screening was based on the titles and abstracts of the records. We did not screen the full texts. If the abstracts of particular items were not displayed properly on the “revtools” Shiny app, the raters would double-check if the abstracts were available using bibliographic databases based on the displayed titles and authors.

### Corpus Scope

The literature search from the four databases resulted in a total of 2167 entries. We removed 805 duplicates automatically, as detected by the “revtools” R package (Westgate, 2019), resulting in 1362 records for screening. After the screening based on titles and abstracts, we excluded 1103 records. The inter-rater reliability (unweighted Cohen’s kappa) of the included 20% of the total number of items (i.e., published articles; *n* = 272) was .592, with a 90.1% agreement rate. Since κ was below .7, Rater 1 went through the complete list of records again based on the consensus reached by both raters and corrected the coding when needed. The full list after screening included 259 records for full-text screening. For the present implementation and demonstration of the MAP-DyS framework, the current version of the corpus focuses on data from papers published between 2014 and 2023. The corpus reflects the coverage of the literature search conducted in December 2022 and captures recent methodological developments in dyslexia subtyping available during the initial phase of framework construction. The resulting dataset comprises 63 studies from 52 items, including early-access publications scheduled for 2023. See the PRISMA flow diagram for details of the study identification and screening processes for constructing this dataset, and the reasons for excluding the screened items (see Appendix).

### Coding Framework and Data Extraction

To operationalise analytic decision pathways across studies, we identified and documented a wide range of variables capturing theoretical choices, sample characteristics, measurement decisions, analytic procedures, and reported outcomes. These variables were pre-defined based on the authors’ domain knowledge and categorised into five key domains: publication information, sample characteristics, measures, methods, and results. Importantly, the variables were refined through discussions among co-authors to ensure comprehensive and meaningful organisation of the extracted data. The final dataset for the app construction consisted of a total of 168 variables. Table 1 outlines some examples of variables in the dataset for illustrative purposes (see also the “List of Variables” tab in the MAP-DyS app for the complete list of variables).

**Table 1.**
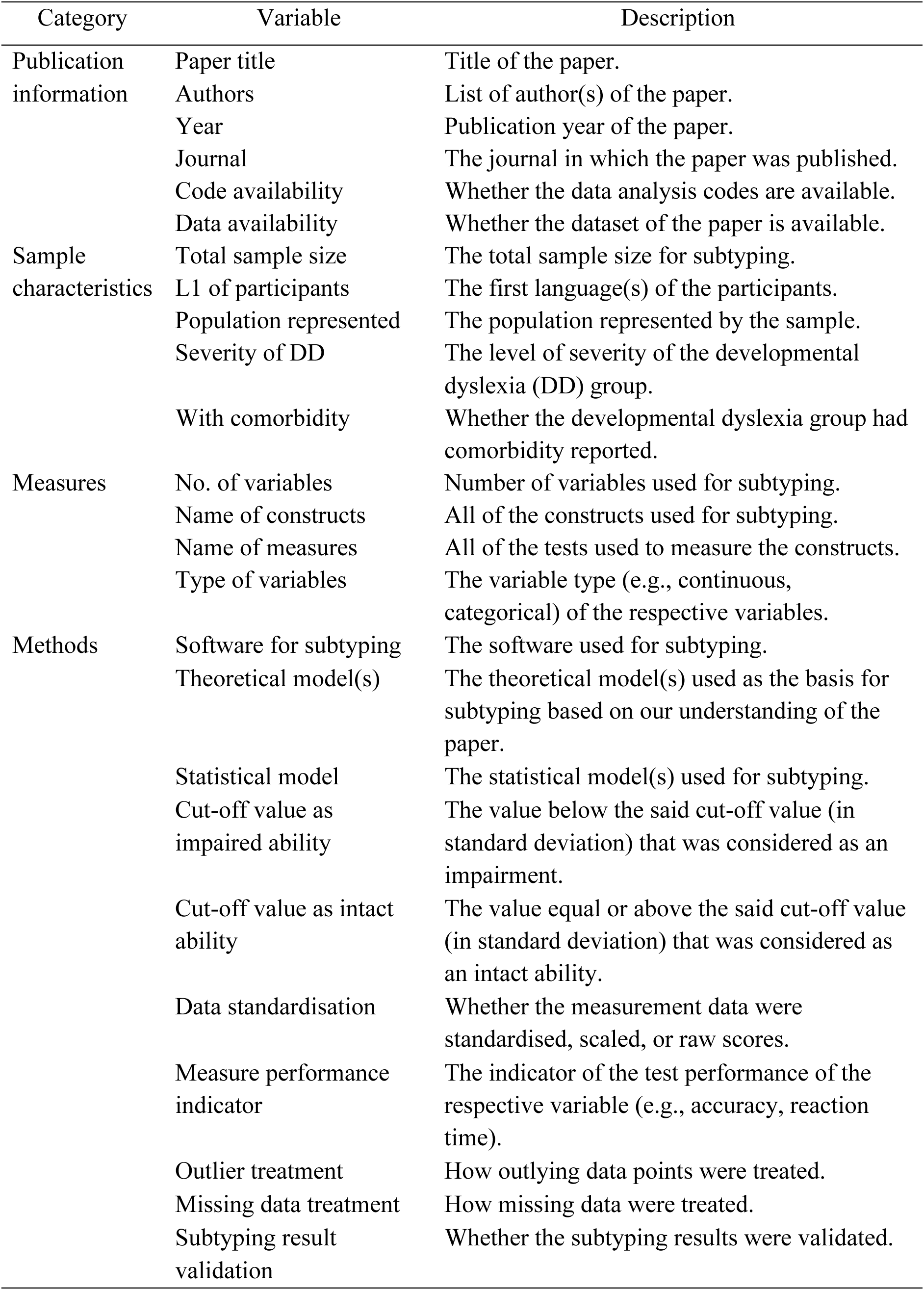
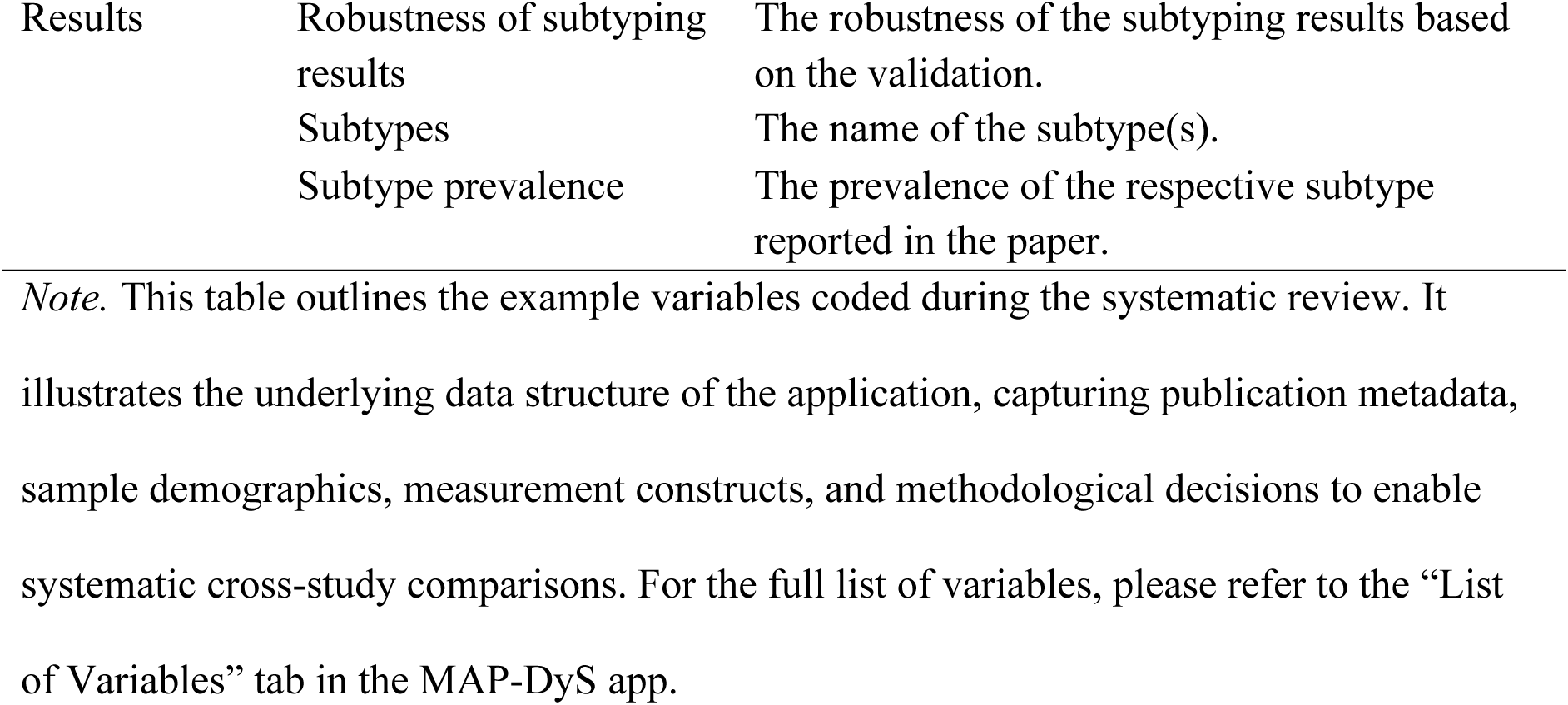
Variables Extracted for the MAP-DyS Dataset.

## MAP-DyS Framework and App Components

### Overview of the Framework

MAP-DyS is an open-source, interactive application designed to support the exploration of analytic decision pathways in dyslexia subtyping research. The hyperlink to the MAP-DyS app can be found via the project GitHub page: https://github.com/kristantodan12/Dyslexia_subtyping. The app consists of three main components: (1) Dataset Exploration to explore and subset the dataset, (2) Multiverse of Decision Steps to investigate the analytic variability, and (3) Subtype Result Explorer to examine the results given specific analytic decisions. At the centre of the homepage is an interactive network map, representing the core components of the app (see Figure 1). Each node corresponds to a specific functionality (see below). When clicking on a node in the network graph, the respective component description will be displayed on the guidance panel on the right. Users can click on the “Go to Tab” button to access the respective component for an in-depth exploration.

**Figure 1.**
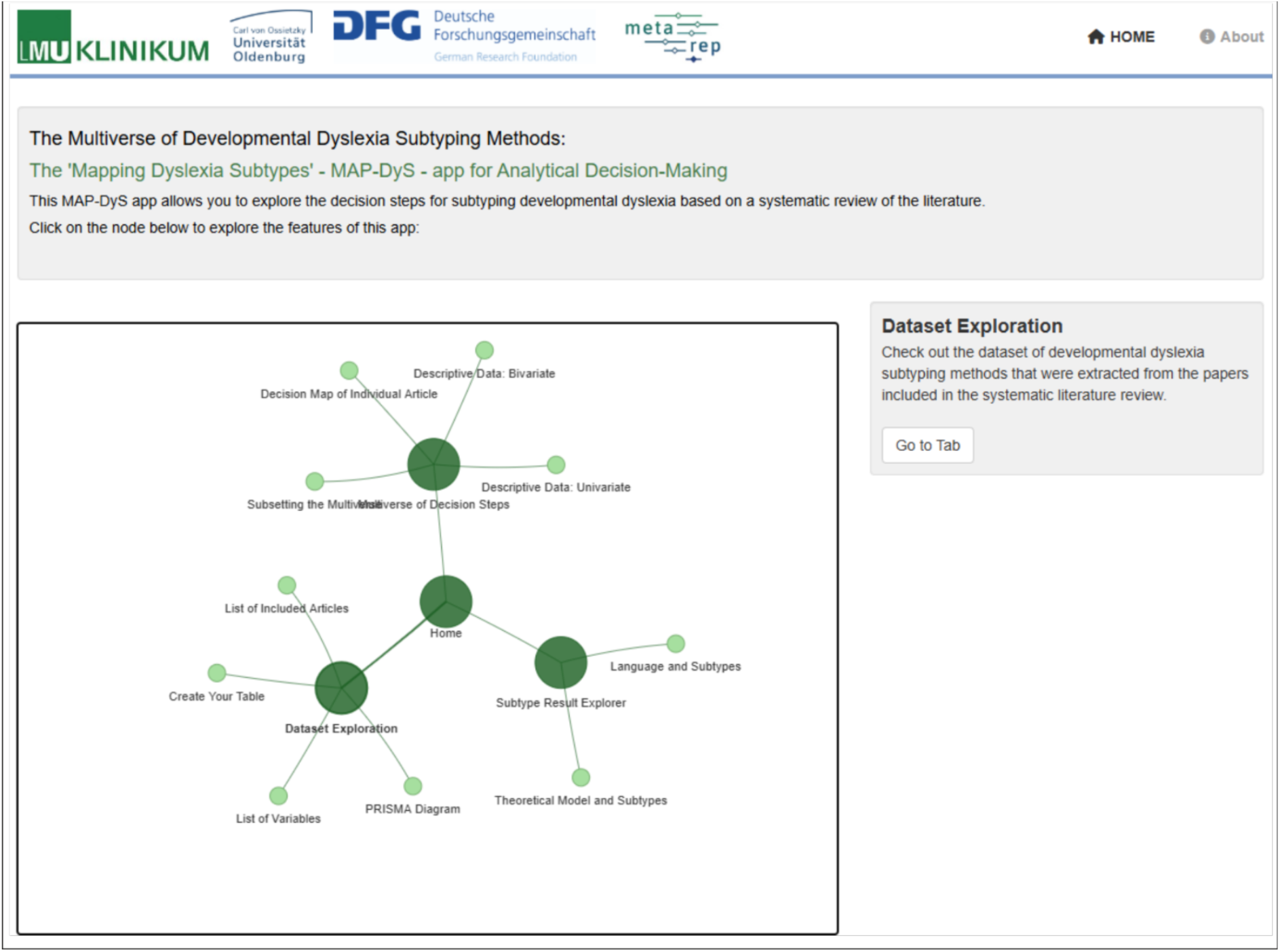
App Component Overview. *Note.* The homepage displays an interactive network map illustrating the core components and functionalities of MAP-DyS. Users can navigate directly to specific data exploration tabs by clicking the corresponding nodes.

### Dataset Exploration

The first component set, “Database Exploration”, consists of four sub-components: PRISMA diagram, list of variables, list of included articles, and create your table. This component set allows users to navigate the general information of the dataset, including a summary of the systematic literature search (“PRISMA Diagram” tab), the list of variables (“List of Variables” tab), and the included articles in the dataset (“List of Included Articles” tab).

A special sub-component is the “Create Your Table” (see Figure 2). This component allows users to generate a tailored subset of the dataset by selecting specific variables of interest. Users can explore the data of the systematic review in a focused and customised manner, catering to their research questions of interest and analytic needs.

**Figure 2.**
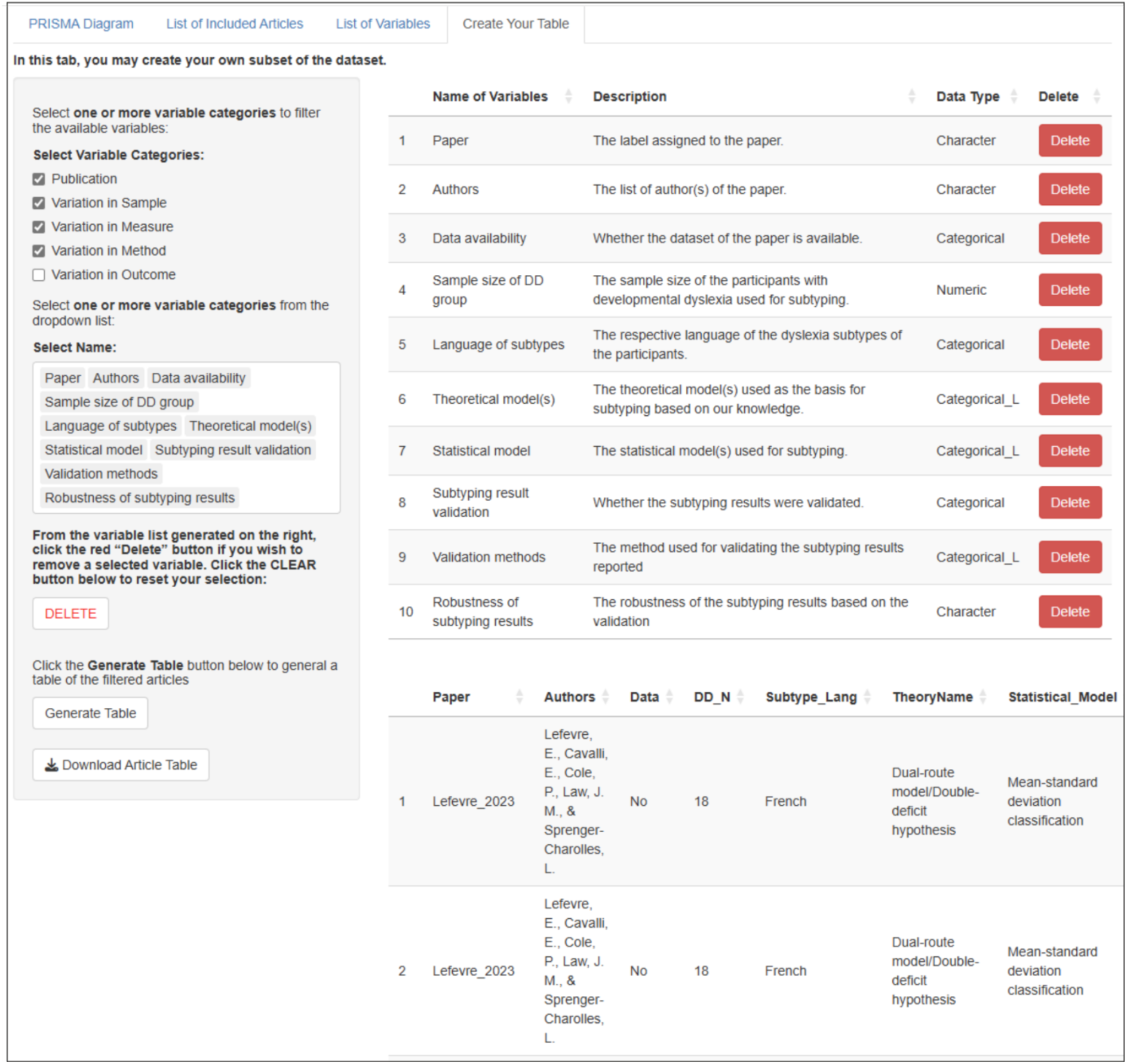
Screenshot of the “Create Your Table” Tab in the “Dataset Exploration” Component Set. *Note.* This interface allows users to build a customised subset of the database. By filtering variable categories on the left sidebar, users can isolate the exact methodological or demographic details relevant to their research. The app then generates a tailored, downloadable data table for focused analysis.

On the left sidebar, users can filter the dataset by selecting one or more variable categories—publication information, sample characteristics, measurement in subtyping, subtyping methodology, and the subtyping outcome. This setup allows users to narrow the scope to the aspects most relevant to a research question. Users can select one or more variables from the filtered dropdown list based on the selected variable category(-ies). The list of selected variables with the variable description and variable type (e.g., character string vs. numerical variables) will then be displayed on the right panel. The unwanted variable(s) can be deleted by clicking the respective “Delete” button at the end of each row of variables. When users click the “Delete” button on the left sidebar, all the selected variables will be deleted. Once the variables of interest are finalised, users can generate a table displaying the filtered subset of the dataset by clicking the “Generate Table” button on the left sidebar. This customised table is immediately displayed below the list of selected variables for further exploration within the app. Users can also click the “Download Article Table” button to download the customised table for offline analyses.

### Multiverse of Decision Steps

The second component set (see Figure 3), “Multiverse of Decision Steps”, focuses on exploring the descriptive data of the variables included in the systematic review (“Descriptive Data: Univariate” and “Descriptive Data: Bivariate” tabs), as well as visualising the multiverse of decision steps involved in dyslexia subtyping (“Subsetting the Multiverse” tab) and the respective decision map in individual articles (“Decision Map of Individual Article” tab).

**Figure 3.**
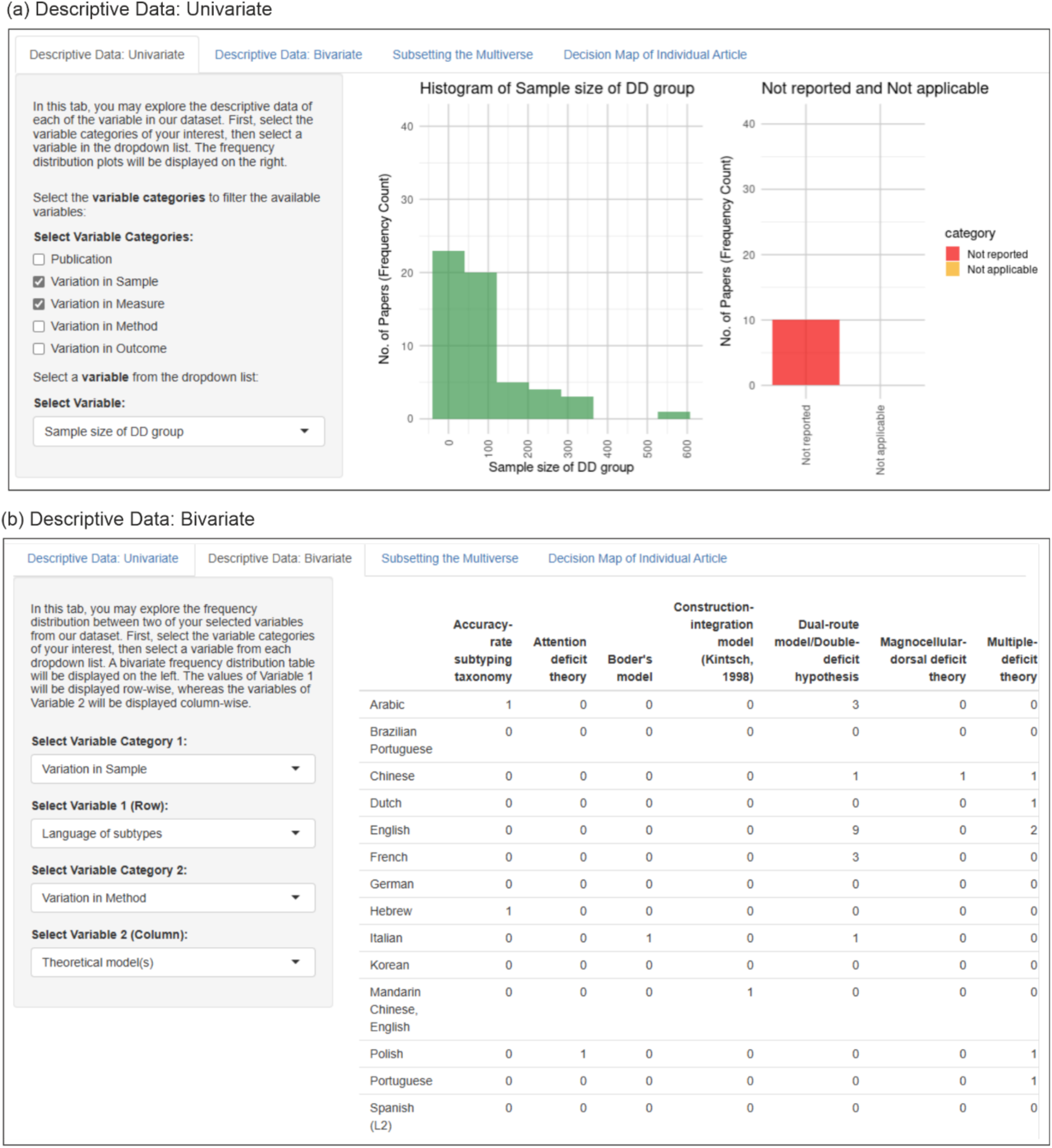
Screenshots of “Descriptive Data: Univariate” Tab (a) and “Descriptive Data: Bivariate” (b) Tab in the “Multiverse of Decision Steps” Component Set. *Note.* Panel (a) maps the frequency distribution of a single variable, such as the sample size of the dyslexic participant group. Panel (b) generates a cross-tabulation matrix to compare two variables simultaneously. As illustrated in panel (b), users can compare the language of the subtypes and the theoretical models used for subtyping reported in studies.

#### Descriptive Data: Univariate

The “Descriptive Data: Univariate” component provides an interactive interface for examining the distribution of individual study-feature variables within the dataset. This allows users to examine the frequency distribution of the variables of interest. Users begin by selecting one or more variable categories to filter the variables available for exploration (e.g., publication year and number of participants in the study). The dropdown menu will display the list of variables, filtered by the chosen variable category(-ies). Once a variable of interest is chosen from the dropdown menu, the component will generate visualisations that display the frequency distribution of the number of reviewed papers across the selected variable in the form of histograms or bar plots. For instance (see Figure 3a), users can select “Sample Size of DD (Developmental Dyslexia) group” under the “Variation in Sample” category on the left sidebar. This produces a histogram illustrating the frequency of sample sizes reported across studies on the right panel. On the far right, a separate bar plot summarises the proportion of papers where sample size data were either not reported or deemed not applicable, represented by distinct red and orange bars.

#### Descriptive Data: Bivariate

The “Descriptive Data: Bivariate” component allows users to explore the relationships between two study characteristic variables from the dataset, providing a matrix-style table that visualises the frequency distribution of two selected variables, enabling the investigation of patterns and associations between the two chosen variables. Users can select any two variables from the dataset to compare. For example, in Figure 3b, the row variable is “Language of Subtypes” (from the “Variation in Sample” category; meaning the first language of the individuals being subtyped), while the column variable is “Theoretical Model(s)” (from the “Variation in Method” category). The resulting matrix table displays the frequency with which different theoretical models are used in studies involving specific participant language groups. With this component, researchers can explore methodological trends, identify research gaps in the literature, and facilitate hypothesis generation.

#### Subsetting the Multiverse

The “Subsetting the Multiverse” component assists in pinpointing articles that meet tailored specifications. Users can begin by selecting a variable category (e.g., Variation in Method, Variation in Sample, etc.) from the dropdown menu. After choosing a specific variable within that category (e.g., “statistical model”), users can select an option associated with the variable (e.g., “K-means clustering”). Clicking the “Add Selection” button adds the specified variable-option pair to a list displayed below. Multiple variable-option pairs can be added sequentially to refine the filtering process further. For example, in Figure 4a, the selected criteria included: (1) language of subtypes: English, (2) name of constructs: phonological awareness, naming speed, and (3) statistical model: k-means clustering. The resulting list of articles that match these criteria will be displayed on the right. The table provides detailed information about the identified studies. Users can remove the most recent selection using the “Delete Last Row” button or start fresh by clearing all selections. This component helps users identify articles with study designs similar to their own, providing useful templates for designing their research.

**Figure 4.**
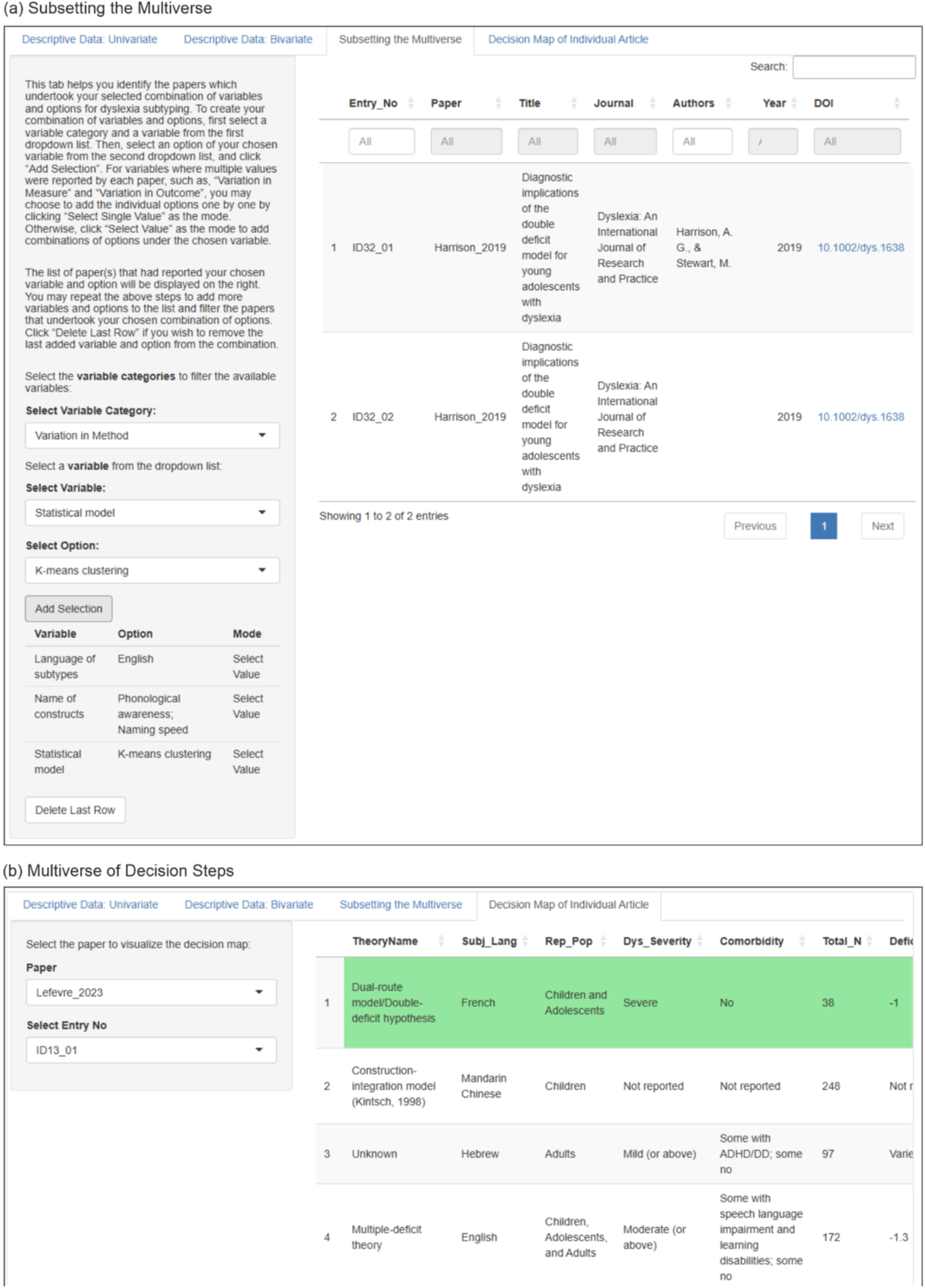
Screenshots of “Subsetting the Multiverse” (a) and “Decision Map of Individual Article” (b) Tabs in the “Multiverse of Decision Steps” Component Set. ***Note.*** Panel (a) shows how users can systematically locate studies that employ the specified analytic pipelines. Panel (b) shows how users can visualise the analytic pipeline of a selected article, highlighted in green, for direct comparisons with the pipelines of other studies in the database.

#### Decision Map of Individual Articles

The “Decision Map of Individual Article” component (Figure 4b) visualises the analytic decisions made in individual studies included in the systematic review. This component allows users to examine specific methodological choices and participant characteristics for each study. At the top of the interface, users can select a specific paper from the “Paper” dropdown menu to view its corresponding decision map. The “Select Entry No.” dropdown menu allows users to select one of the reported entries of results from the selected paper in case there is more than one entry of results. Once a paper is selected, a table summarising its decision steps is displayed on the right panel. Each column represents a decision step, such as the theoretical model applied, the language of participants, and population characteristics. Each row represents a study. The displayed rows represent studies that share one or more analytic decisions with the selected paper. If a study (represented by a row) has an overlapping decision at certain steps with the selected paper, that decision step will be highlighted in green. Users can identify patterns, inconsistencies, and areas of overlapping decisions among studies.

### Subtyping Result Explorer

#### Theoretical Model and Subtypes

The “Theoretical Model and Subtypes” component is for examining the relationships between theoretical models and specific dyslexia subtypes included in the systematic review (see Figure 5a). The central component is a heatmap, where each cell represents the number of data entries associated with a specific subtype (e.g., phonological dyslexia, surface dyslexia, double-deficit) under a particular theoretical model (e.g., dual-route model, multiple-deficit theory). The dyslexia subtype names on the X-axis are grouped by their categories (i.e., cognitive subtype vs. behavioural subtype), indicated by the blue category label preceding each subtype name. The colour intensity of each cell reflects the frequency of data entries, with darker shades indicating higher counts. Users can hover over any cell to view a tooltip displaying detailed information about the corresponding subtype, theoretical model, and the number of data entries associated with the respective cell.

**Figure 5.**
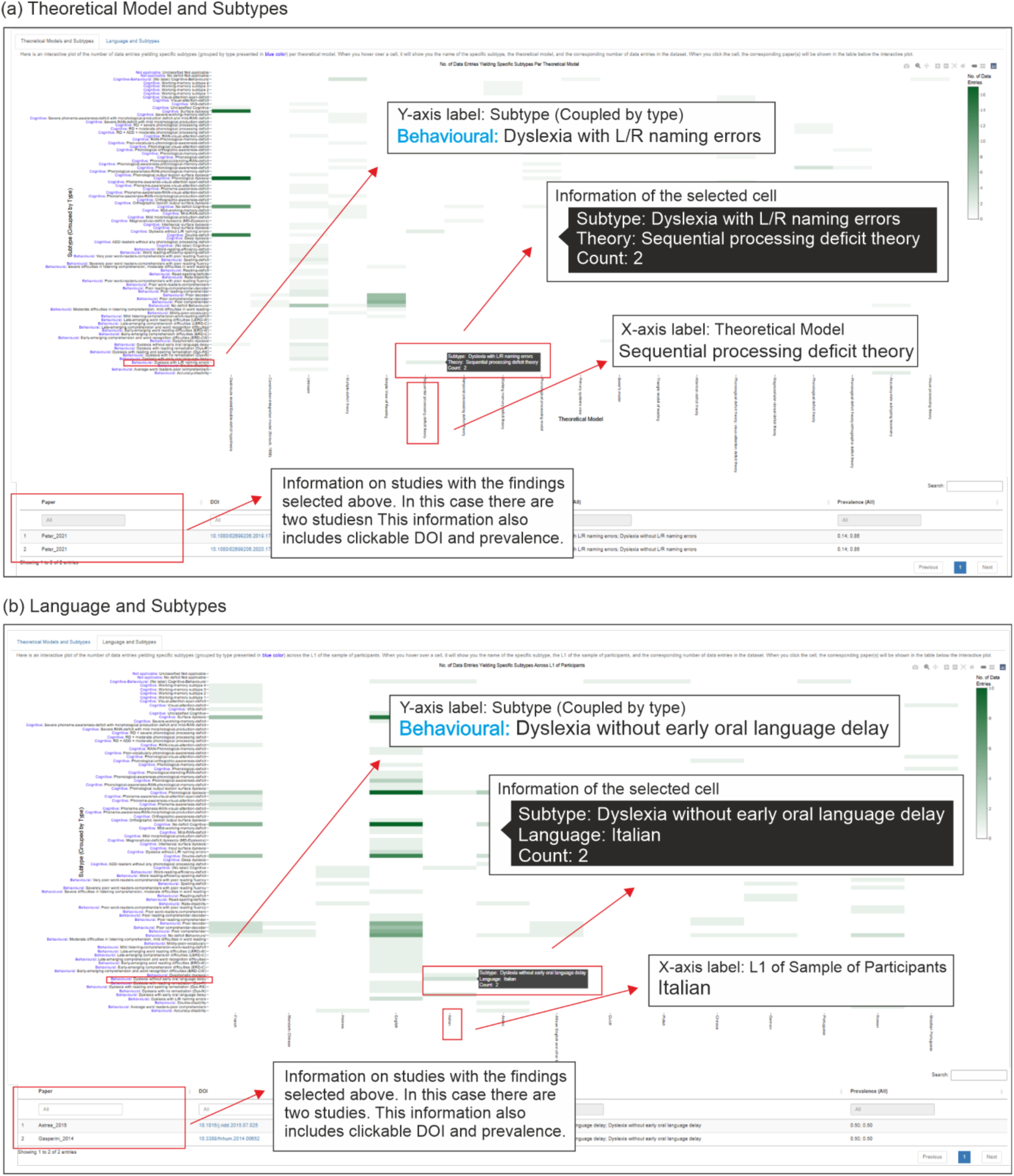
Screenshots of the “Theoretical Model and Subtypes” (a) and “Language and Subtypes” (b) Tabs in the “Subtyping Result Explorer” Component Set. *Note.* Panel (a) is a heatmap that shows how specific dyslexia subtypes emerge from various theoretical frameworks. Panel (b) is a heatmap that shows the distribution of identified subtypes across different participant languages. Darker cells indicate higher frequencies, and clicking on any cell retrieves a sortable table of the specific studies contributing to that methodological intersection.

To delve deeper, users can click on a specific cell on the heatmap. Upon selection, a dynamic table appears below the heatmap, listing the studies contributing to that specific combination of theoretical model and subtype. This table provides the key information, including the study title, theoretical framework, identified subtypes, and prevalence rates of the studies displayed. Users can search, filter, and sort the table to focus on studies of interest, facilitating targeted exploration. This feature allows users to trace the theoretical foundations and methodological decisions behind specific subtype classifications in the dataset.

#### Language and Subtypes

The “Language and Subtypes” component (see Figure 5b) visualises the relationships between the first language(s) of participants and the subtypes of dyslexia identified in the systematic review. This component contains a heatmap, where each cell represents the number of data entries linking a particular language (e.g., French, English, Mandarin) to a specific dyslexia subtype (e.g., phonological dyslexia, surface dyslexia, double-deficit). Similar to the “Theoretical Models and Subtypes” component (Figure 5a), the subtype names on the X-axis are grouped according to their subtype categories (cognitive subtype vs. behavioural subtype; indicated by the blue words). The intensity of the cell colour corresponds to the frequency of data entries, with darker shades indicating a higher count. Hovering over a cell reveals a tooltip displaying the subtype, the language, and the count of associated data entries. For example, in the provided screenshot, the intersection of “surface dyslexia” and “French” indicates four data entries.

Clicking on a cell dynamically generates a table below the heatmap, listing the studies corresponding to the selected language-subtype combination. The table provides detailed information, including the study title, DOI, theoretical framework, subtypes identified, and prevalence rates of each identified subtype within the corresponding study. This function allows users to trace the studies contributing to the observed patterns and examine their methodological details.

## Demonstration of the MAP-DyS Framework Using Dyslexia Subtyping Studies

To demonstrate the utility of MAP-DyS, we present illustrative outputs in three areas: variability in metadata and sample characteristics, variability in analytic decision steps, and variability in reported subtyping results. The figures in this section are captured from the MAP-DyS interface. Readers are encouraged to use the app in parallel to explore additional patterns beyond the examples shown here.

### Variability in Metadata and Sample Characteristics

We first evaluate the variability in the study metadata and sample characteristics. As depicted in Figure 6a, the distribution of publications reveals temporal variation across the sampled period (2014–2023), with relatively higher representation in the earlier and later years, reflecting an increase in the number of available studies over time. This pattern may reflect broader shifts in theoretical and methodological approaches within the field, including periods of critical reflection on the heterogeneous nature of dyslexia (see the “Dyslexia Debate”; Elliott & Grigorenko, 2014, 2024; Kirby, 2020; Peterson & Pennington, 2012; Ramus, 2014) and more recent uptake of data-driven techniques (Feczko et al., 2019). While these explanations regarding the number of publications are not directly tested here, they suggest that subtyping practices are embedded within changing research contexts. Using MAP-DyS, researchers can examine how analytic decisions cluster within different time periods—for example, identifying whether certain statistical approaches or theoretical frameworks are more prevalent in recent versus earlier studies—and use this information to make more informed and context-aware methodological choices.

**Figure 6.**
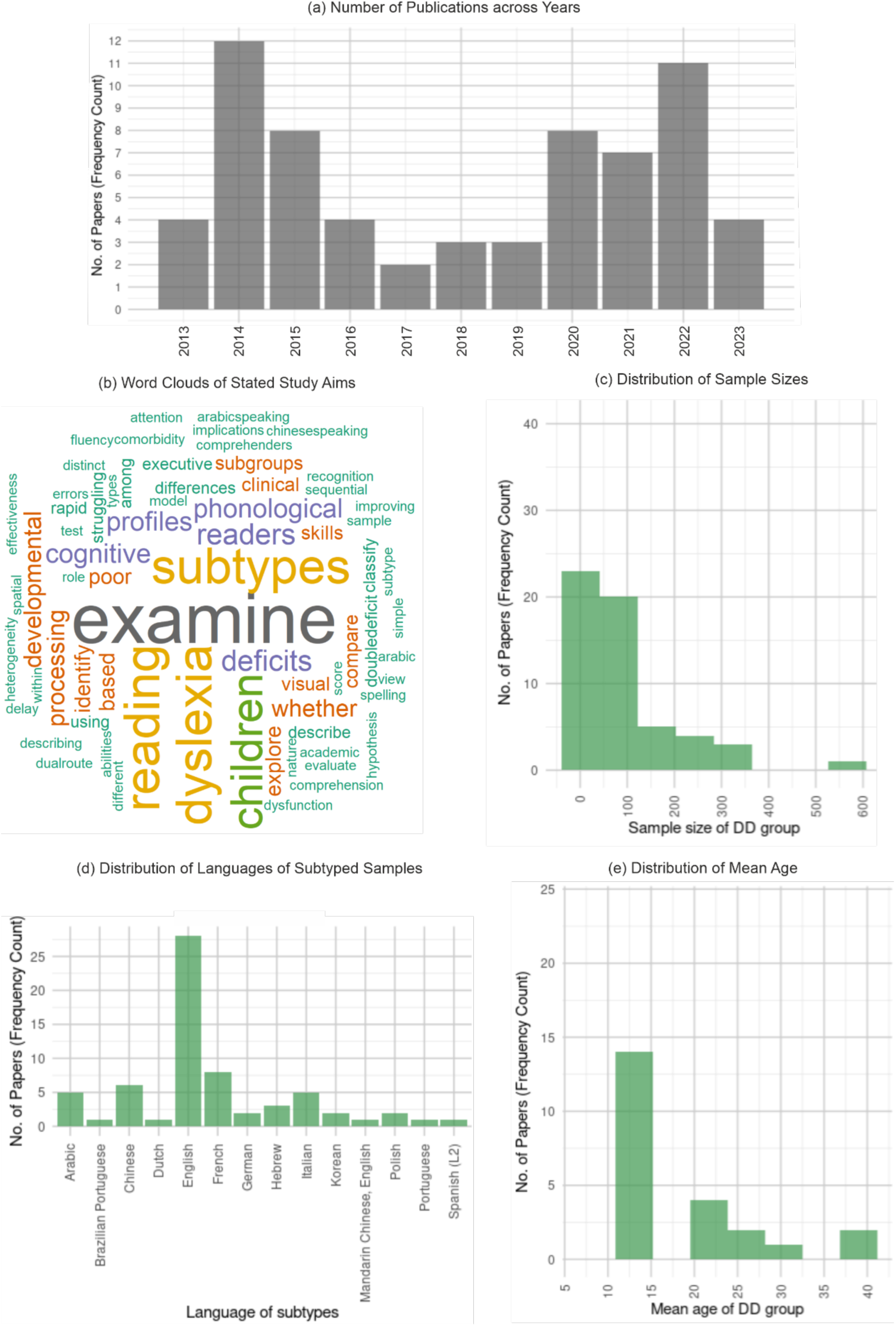
Empirical Overview of Dyslexia Subtyping Studies: Study Distribution, Research Focus, and Sample Characteristics.

We next consider the stated aims of the included studies. Figure 6b visualises these aims via a word cloud, which underscores the descriptive and taxonomic focus of the field. The prominent terms include “examine”, “identify”, “profiles”, and “subtypes”, alongside theoretical constructs such as “phonological” and “cognitive”. This pattern suggests that the literature has primarily emphasised the identification and description of heterogeneity. By examining these patterns in MAP-DyS, researchers can reflect on how study aims align with analytic strategies—for instance, distinguishing between exploratory subgroup identification and studies that incorporate validation or predictive modelling, and use this information to more deliberately define the scope of their own analyses.

Finally, with regard to sample characteristics, Figures 6c–6e reveal imbalances in sample size, language, and participant age. Sample sizes are predominantly small, with most studies relying on fewer than 100 participants to derive subtypes. While mapping this information allows researchers to benchmark their own designs and recognise the risks to statistical stability, it also highlights a clear need for larger datasets. One potential avenue for overcoming these data limitations is the aggregation of data across studies. Specifically, approaches such as federated learning enable models to be trained collaboratively across distributed datasets without requiring direct data sharing (Rieke et al., 2020). By allowing models to be trained across distributed datasets without direct data sharing, federated learning offers a practical solution to the small-sample bottlenecks that frequently compromise model estimation in current subtyping research.

Beyond the sheer size of the samples, our mapping reveals that the literature is also constrained by a lack of demographic and linguistic diversity. Geographically and linguistically, the literature is heavily skewed toward Western contexts: English is the most frequently studied language, followed by French, while other languages (e.g., Chinese, Arabic, Korean) are less represented. Mapping these distributions enables researchers to assess the generalisability of subtype structures across linguistic contexts and to identify underrepresented populations. In terms of developmental focus, the mean age of participants clusters around early adolescence (11–15 years), with fewer studies examining early childhood or adult populations. This highlights opportunities for more balanced sampling across the lifespan.

### Variability in Analytic Decision Steps

#### Theoretical Frameworks and Construct Selection

To examine variability across the subtyping pipeline, we analysed how studies differ in a series of analytic decisions, including the selection of theoretical frameworks and the operationalisation of constructs used for subtype identification. The MAP-DyS dataset includes a broad set of variables capturing multiple stages of the subtyping pipeline (see Table 1). In the present section, we focus on analytically consequential decision points that are particularly informative for illustrating variability in analytic choices.

Theoretical frameworks varied widely across studies (Figure 7a). The Dual-Route Model of reading/Double-Deficit Hypothesis was the most frequently cited framework (e.g., Coltheart, 2005), while a substantial proportion of studies (*N* = 13, ∼20%) did not explicitly specify a theoretical model (“Unknown”). The Simple View of Reading emerged as the second most frequently specified framework (Hoover & Gough, 1990). These patterns indicate that both theory-driven and more exploratory approaches coexist in the literature, suggesting that subtyping practices and the resulting subgroup structures may differ. This depends on whether they are guided by explicit theoretical assumptions or more data-driven strategies.

**Figure 7.**
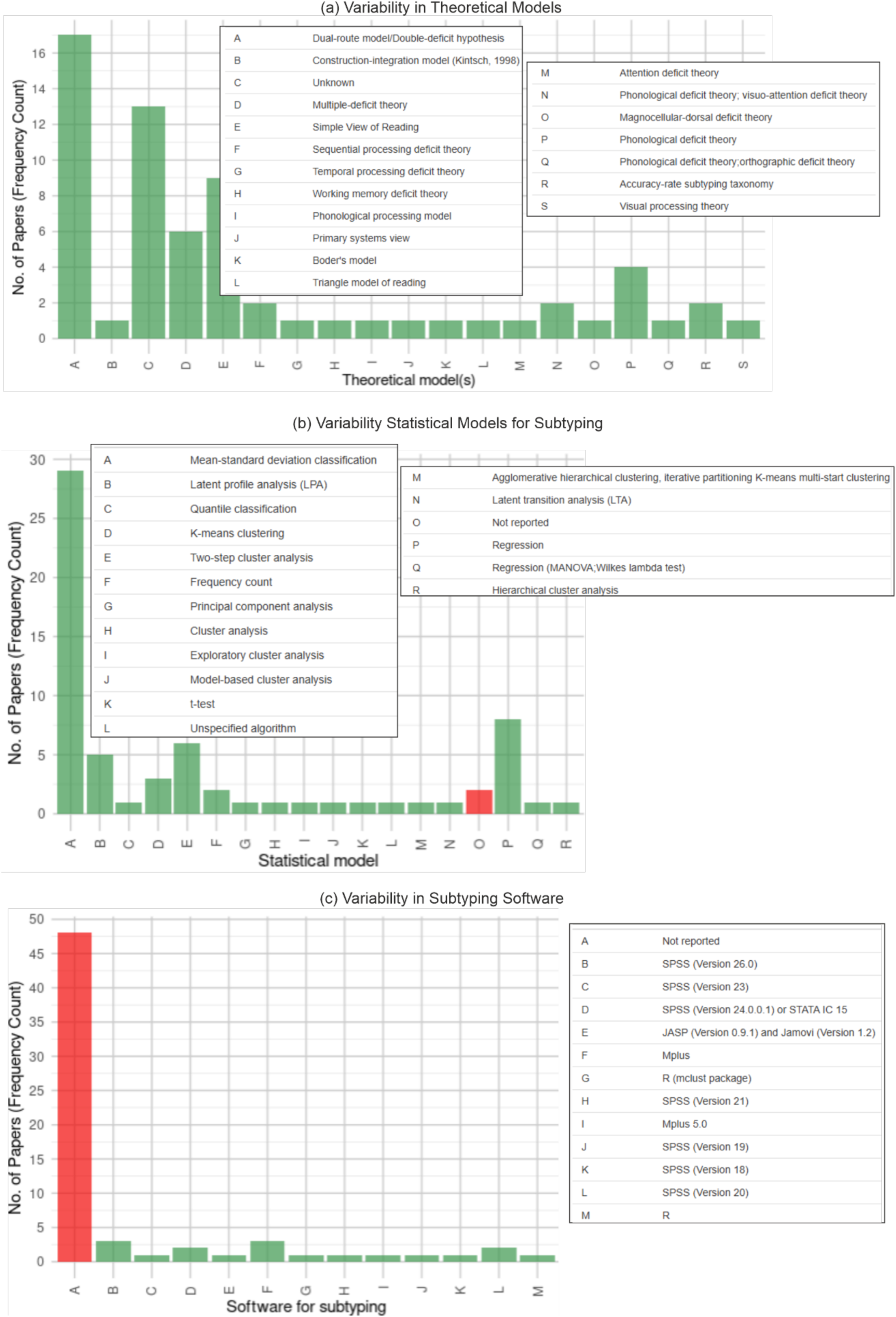
Methodological Variability in Dyslexia Subtyping: Software, Theoretical Models, and Statistical Approaches.

Consistent with this variability in theoretical grounding, we observed substantial differences in construct selection and operationalisation. Different theoretical frameworks prioritise different dimensions of reading and language (e.g., phonological processing, decoding, or language comprehension), which in turn shape the constructs selected for subtyping. At the level of measurement, variability was evident not only in the domains included (i.e., the cognitive or linguistic abilities assessed, such as reading, language comprehension, or memory), but also in the terminology used to describe conceptually related but not necessarily equivalent constructs. For instance, different labels were used for reading-related abilities, such as word identification, word recognition, and real-word reading, as well as for non-word processing (e.g., pseudoword processing, phonological decoding). Importantly, we do not assume that these labels refer to identical constructs, as their interpretation may depend on the underlying theoretical framework and the level of analysis (e.g., broad constructs versus more specific subcomponents).

In addition, the composition of test batteries (i.e., the combination of measures used as input for subtype identification) varied widely across studies. The most frequent combination, language comprehension paired with decoding, was used in only four of the included studies, highlighting a lack of consensus regarding which dimensions should be included. As these measurement choices define the feature space on which subtyping is performed, they represent a critical source of variability in downstream results. Given the heterogeneity in terminology and the absence of a one-to-one correspondence between labels and constructs, this variability is not readily reducible to a single summary figure. Instead, MAP-DyS allows researchers to explore these decisions interactively, for example, by examining how different theoretical frameworks are associated with specific construct selections across studies.

#### Statistical Modelling Approaches

Methodological choices for subtype identification revealed substantial variability (Figure 7b). The most prevalent method was mean-standard deviation classification (*N* = 29, ∼46%), in which subtypes are defined by applying pre-set clinical cut-offs to continuous scores (e.g., Shany et al., 2022). This approach reflects a traditional, clinically oriented framework in which categorical distinctions are imposed on continuous dimensions.

In contrast, data-driven methods were more heterogeneous and less consistently applied. Regression-based discrepancy models (*N* = 8, ∼13%), two-step cluster analysis (*N* = 6, ∼10%), and Latent Profile Analysis (LPA) (*N* = 5, ∼8%) represented the primary alternatives. These approaches differ in their assumptions about subgroup structure, ranging from predefined discrepancy criteria to probabilistic latent class estimation. Notably, two studies failed to report their statistical algorithm entirely, further highlighting gaps in methodological transparency.

The observed heterogeneity in statistical approaches also highlights a broader shift toward data-driven methods in subtyping research. While traditional approaches rely on predefined theoretical assumptions, more flexible clustering and machine learning techniques offer the potential to identify subgroup structures without strong a priori constraints. However, such approaches are highly dependent on data quality, sample size, and validation procedures, and their application in the current literature remains uneven.

#### Validation Practices

Importantly, our review identified a lack of robust validation procedures. More than half of the included studies did not report any formal validation of their identified subtypes (e.g., robustness checks or external validity). Among the studies that did perform validation, methods typically included split-half cross-validation or comparison with alternative clustering solutions (robustness checks). While these validated studies generally reported consistent subtype structures, the high prevalence of unvalidated findings raises questions about the generalisability of the broader literature. Readers are encouraged to explore the specific validation strategies and their outcomes using the Variation in Method > Subtyping result validation filter in the MAP-DyS app.

#### Software and Computational Environment

Transparency regarding the computational tools used for subtyping was notably poor (Figure 7c). The majority of studies (*N* = 48, ∼76%) did not report the specific software or version used for their analysis, creating a barrier to reproducibility. Among the studies that did report their tools, SPSS (across multiple versions) dominated the landscape (*N* = 10, ∼16%), while open-source, script-based environments such as R were used in only one case. This pattern has direct implications for reproducibility. The limited reporting of software, combined with the predominant use of closed-source environments, constrains the transparency of analytic workflows and the ability to reproduce results. In contrast, open-source, script-based environments (e.g., R or Python) require researchers to explicitly document every analytic step in code. This generates a transparent, executable record of the entire pipeline, allowing independent researchers to replicate the subtyping results.

### Variability in Subtyping Results

The final output of the subtyping pipeline, the number and nature of the identified subgroups, exhibited substantial variability. Consistent with earlier sections, this variability appears to be shaped by upstream analytic decisions, including theoretical frameworks, construct selection, statistical modelling choices, and sample characteristics. The MAP-DyS dataset captures a wide range of outcome variables (Table 1). Here, we focus on key patterns in subgroup structure that illustrate how different analytic pathways lead to different subtyping results.

Regarding the granularity of classification, most studies reported relatively low numbers of subtypes. As illustrated in Figure 8, the majority of studies identified between two and four subtypes, with solutions exceeding four subgroups being comparatively rare. This pattern likely arises from a combination of constraints and assumptions across the subtyping pipeline. One contributing factor may be statistical constraints, as many studies relied on small sample sizes (*n* < 100), which may restrict the stability of more complex solutions. In addition, theoretical frameworks may implicitly constrain the number of expected subtypes; for example, models such as the “dual-route” (Coltheart, 2005), or “simple view of reading” (Hoover & Gough, 1990), tend to foreground a limited set of core dimensions, which naturally yield a small number of subtypes. Finally, simpler solutions may also be favoured in practice due to their interpretability and clinical usability, as more granular classifications are often harder to validate and apply (Kolyshkina & Simoff, 2021).

**Figure 8.**
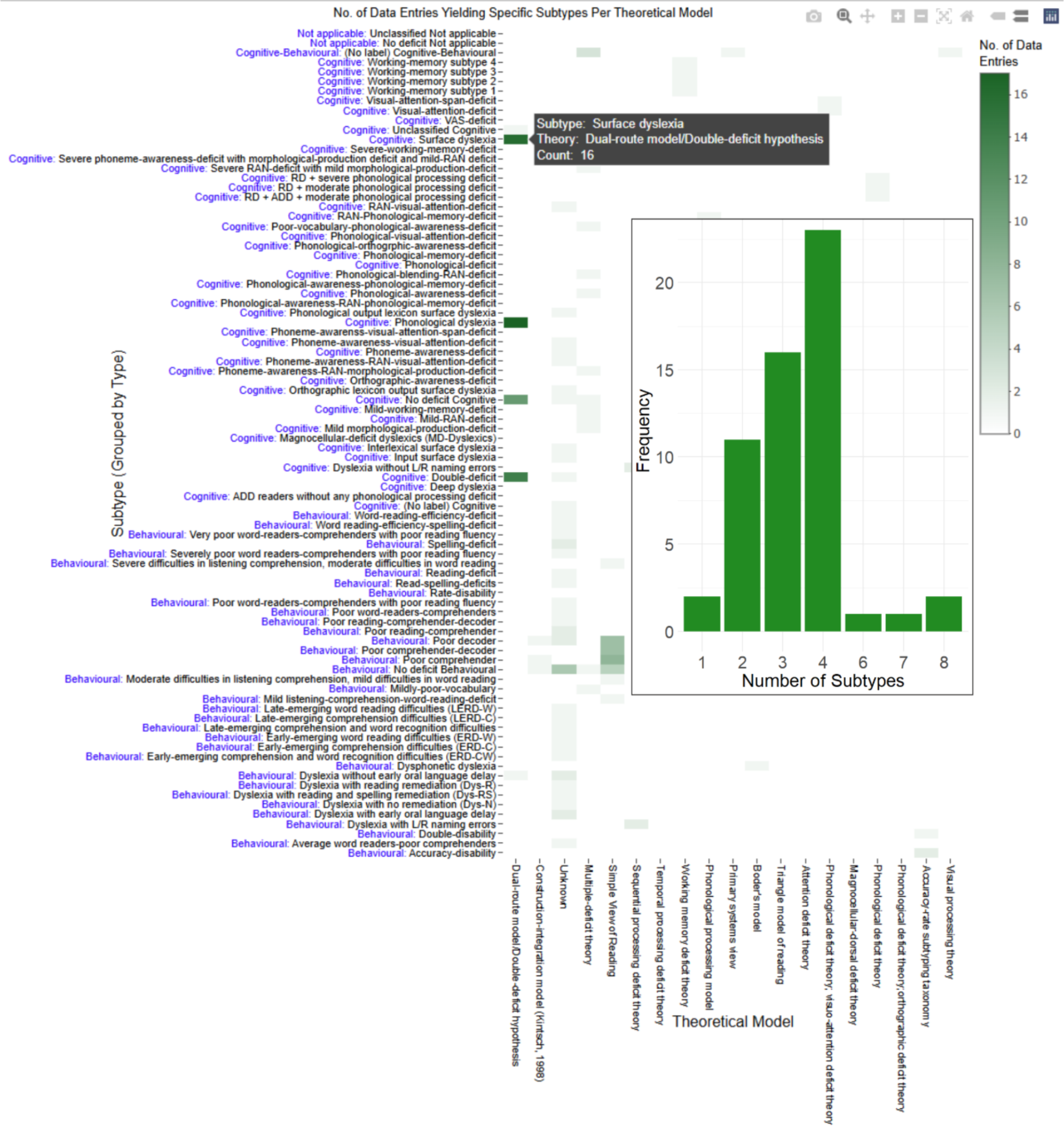
Distribution of Identified Subtypes Across Theoretical Frameworks. *Note.* The heatmap displays the frequency with which specific dyslexia subtypes were reported across theoretical frameworks in the included studies. The inset histogram shows the distribution of the total number of subtypes identified per study. Frequencies reflect the number of extracted data entries.

Moreover, using the Subtype Result Explorer within the MAP-DyS app, we mapped the relationship between participants’ first languages and the identified subtypes. While certain profiles, such as phonological and surface dyslexia (and their combination), appeared relatively consistent across languages (including English, French, Arabic, and Chinese), additional language-specific profiles also emerged. These included working memory subtypes in French samples, combined rapid automatised naming (RAN) and morphological production deficits in Chinese, and average word readers with poor comprehension in Korean. However, given the limited number of studies in several language groups, these differences cannot be straightforwardly attributed to language-specific cognitive profiles, as they may also reflect differences in theoretical frameworks, construct selection, and measurement practices across studies. For researchers, this highlights a key implication: subtype labels observed in a given language should not be interpreted in isolation, but in relation to the analytic decisions that produced them. The MAP-DyS app enables this by allowing users to trace how language, theoretical assumptions, and measurement choices co-occur with specific subtype solutions, thereby supporting more informed interpretation and comparison of subtyping results across studies.

This relatively limited number of subtypes, as reflected in our dataset, is noteworthy, as it contrasts with theoretical accounts that propose a larger set of distinct dyslexia profiles (e.g., Zoccolotti & Friedmann, 2010). This discrepancy raises the possibility that the number of identified subtypes may be shaped not only by theoretical assumptions, but also by methodological constraints such as sample size, variable selection, and modelling choices. Distinguishing between these influences requires designs that systematically vary analytic decisions within the same dataset.

## General Discussion

### Implications for Research and Practice

By visualising how subtypes vary according to theoretical models, participant populations, and statistical techniques, the MAP-DyS framework shifts how subtyping research can be understood and evaluated. Rather than treating subtypes as fixed entities, our app makes explicit that subtyping outcomes are contingent on a series of analytic decisions. This enables researchers to examine subtyping results not only at the level of outcomes, but also at the level of the methodological pipelines that produce them.

One key contribution of the MAP-DyS app is that it facilitates comparative analyses between analytic decisions and the corresponding outcomes across dyslexia subtyping studies. The app systematically maps how different methodological choices are distributed across studies. This structured overview enables researchers to generate hypotheses about how specific combinations of theoretical frameworks, constructs, and statistical models are associated with particular subtype solutions, thereby reframing subtyping as a decision-dependent process rather than a purely discovery-driven one.

Crucially, our framework highlights the extent of methodological variability within subtyping research and makes this variability inspectable. For instance, even when studies adopt the same statistical technique, such as cluster analysis (Harrison & Stewart, 2019; Houpt et al., 2015), they may differ substantially in key decision steps, including how optimal solutions are selected and how results are validated. The explicit visualisation of these methodological pipelines encourages researchers to document methodological decisions and outcomes clearly, so as to enhance research reproducibility through such transparency (Weissgerber et al., 2016). It also allows researchers to discern reasonable decisions from arbitrary ones and assess whether these methodological choices are theoretically and methodologically justified (Del Giudice & Gangestad, 2021; Olsson-Collentine et al., 2023; Parsons, 2022; Steegen et al., 2016).

Importantly, our framework also reveals that the existing literature predominantly focuses on identifying subtypes, while comparatively less attention is devoted to validation and implementation. By surfacing this pattern, MAP-DyS enables researchers to critically assess not only how subtypes are derived, but also whether they are supported by sufficient validation procedures and whether they are positioned for meaningful application. This shifts the evaluation of subtyping research from a focus on classification alone to a broader consideration of robustness and utility.

Beyond research applications, our interactive app may provide practical value for clinicians, educators, and policymakers by presenting a structured overview of how subtypes have been defined and identified across studies and populations. While not a diagnostic tool, it can support evidence-informed reflection on diagnostic and intervention practices (Mather & Wendling, 2024). Clinicians may consider the most consistently applied subtyping frameworks as one reference point for aligning subtyping methods and intervention practices. Additionally, educators can use the app to explore cognitive subtypes identified in research, supporting a better understanding of learner diversity and informing instructional strategies.

From a broader perspective, the MAP-DyS framework contributes to ongoing meta-scientific efforts to understand and document methodological variability in psychological research (Stanley et al., 2018). By making analytic decision pathways explicit, the framework encourages a shift from viewing variability as noise to recognising it as an inherent feature of complex research pipelines. This perspective promotes more cautious interpretation of heterogeneous findings and supports efforts to enhance the replicability and generalisability of research (Usui et al., 2021).

Furthermore, our framework provides a basis for comparing subtyping practices across domains. For instance, our app enables researchers to examine how dyslexia subtypes are defined and validated, and to compare these practices with subtyping approaches in fields such as autism spectrum disorder (Agelink van Rentergem et al., 2021), where more established validation protocols may offer useful reference points. In this way, MAP-DyS functions as an interactive meta-research tool that supports cross-field methodological reflection and synthesis. In line with recommendations from clinical research (Gagnier et al., 2012, 2013), our framework reinforces the importance of transparent, systematic, and comprehensive reporting of analytic decisions to enable meaningful comparison and synthesis across studies.

Finally, by making analytic variability explicit and explorable, MAP-DyS provides a foundation for multiverse-aware research. Researchers can use the app to identify alternative analytic pathways, assess how common they are in the literature, and design robustness checks or multiverse analyses accordingly. In doing so, our framework supports a shift toward more transparent and systematically evaluated analytic practices in subtyping research.

### Limitations and Future Directions

While the app provides an innovative and systematic approach to exploring the multiverse of dyslexia subtyping methods, several limitations remain. The current dataset relies on recently published studies, which may introduce biases due to incomplete reporting and variability in study designs. Researchers may have explored additional options beyond those they report in their published work. The utility of the dataset also depends on the quality and granularity of the extracted data. Missing information in the original studies (e.g., preprocessing methods, subgroup validation, or exact statistical thresholds) may limit the extent to which users of the app can compare analytic decisions and research pipelines across studies. Furthermore, the app currently focuses on predefined decision steps, which, while extensive, may not capture emerging methodological developments or alternative approaches not widely adopted in existing research. Thus, some decision steps and options may not yet be extracted and reported in our dataset. Notably, we are actively planning to expand the dataset with publications from earlier years, which will be integrated in subsequent releases of the tool. Future research could extend our work by performing full multiverse analyses on individual studies to examine how results vary depending on analytic choices, a direction that highlights the importance of open data and reproducible pipelines in subtyping research.

Beyond dyslexia research, the app’s framework could be adapted to other fields grappling with methodological variability in subtyping. Many disciplines within psychology, cognitive science, and biomedicine, such as autism spectrum disorders (Mottron & Bzdok, 2020), ADHD (Karalunas & Nigg, 2020), and psychiatric disorders like schizophrenia (Joyce & Roiser, 2007), face similar methodological challenges in subtyping, so as to understand the cognitive heterogeneity of these constructs (Feczko et al., 2019). Our app’s publicly available code provides a flexible, reproducible tool that can be tailored to these contexts. By applying this multiverse visualisation approach, researchers across disciplines can systematically explore subtyping decisions to better understand the heterogeneity of complex constructs.

## Conclusion

Our interactive MAP-DyS app provides a systematic framework for visualising and interrogating methodological variability in subtyping research. By demonstrating that subtyping outcomes are shaped by analytic decision pathways, our work reframes how subtyping results can be interpreted, evaluated, and compared across studies. Rather than offering a single subtyping solution, our framework enables researchers to navigate the complexity of subtyping research more transparently and critically.

Beyond dyslexia, the MAP-DyS framework offers a generalisable approach for examining methodological variability across psychological and biomedical domains. Moving forward, we encourage continued collaboration within the research community to refine and expand the framework and app. This ongoing development would support more transparent, reproducible, and multiverse-aware analytic practices, contributing to ongoing efforts to improve research quality and interpretability in behavioural science.

## Acknowledgements

We thank Peter Roßmeier for assisting in the first stage of coding in the systematic literature review.

## Data and Code Availability

The datasets and R scripts are publicly available in our GitHub repository: https://github.com/kristantodan12/Dyslexia_subtyping

## Ethics Approval

Not applicable.

## Authors’ Contributions

**Anna Yi Leung:** Conceptualisation, Data curation, Formal analysis, Funding acquisition, Investigation, Methodology, Project administration, Software, Resources, Validation, Writing—original draft, Writing—review & editing. **Daniel Kristanto:** Conceptualisation, Formal analysis, Funding acquisition, Investigation, Methodology, Project administration, Software, Resources, Validation, Visualisation, Writing—review & editing. **Carsten Gießing:** Methodology, Supervision, Validation, Writing—review & editing. **John PA Ioannidis:** Methodology, Supervision, Validation, Writing—review & editing. **Andrea Hildebrandt:** Methodology, Supervision, Validation, Writing—review & editing. **Xenia Schmalz:** Methodology, Supervision, Validation, Writing—review & editing.

## Appendix

### PRISMA Flowchart Diagram of Study Identification

This diagram outlines the systematic literature search and screening process used to construct the MAP-DyS dataset.

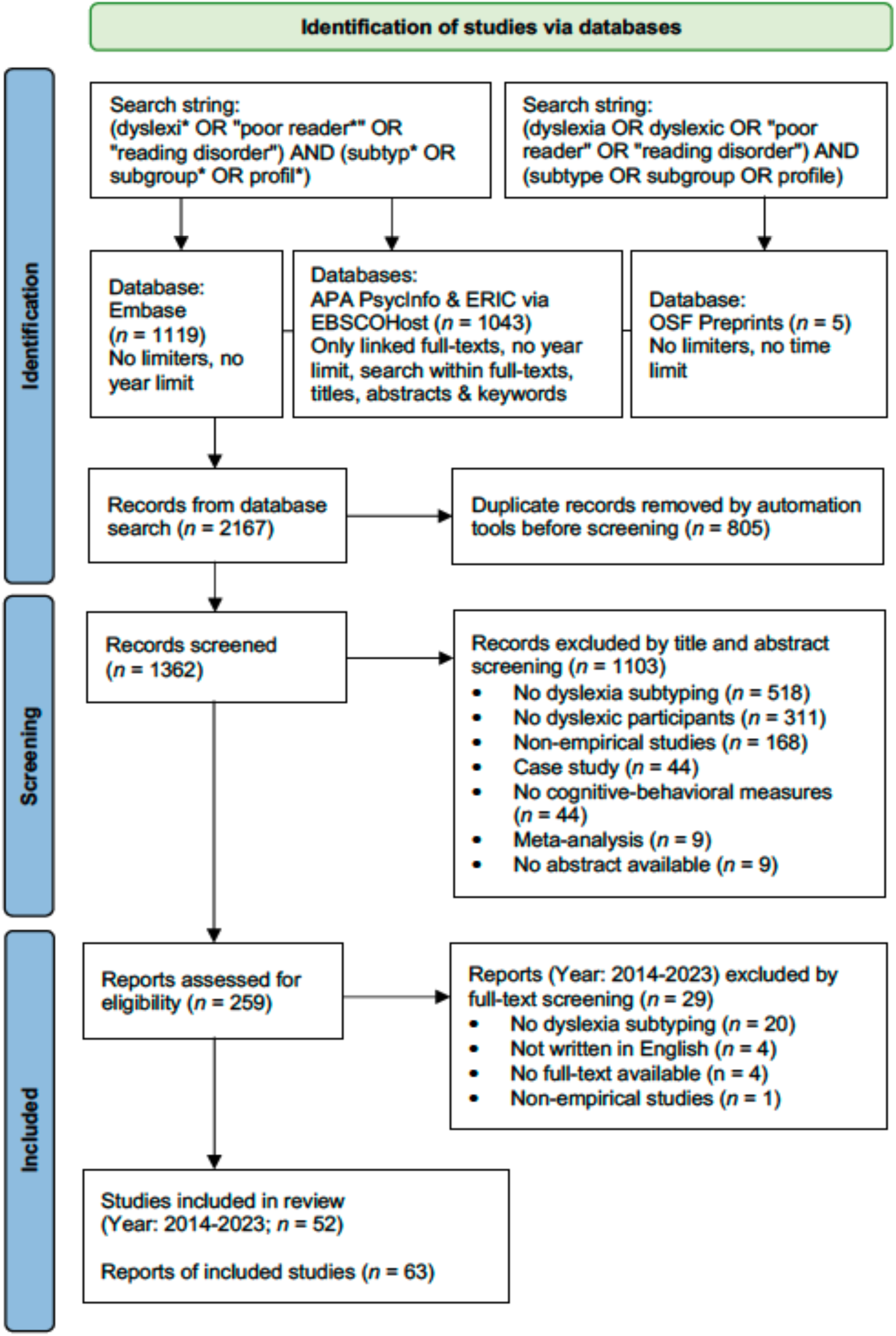

